# DETERMINANTS OF COMPLIANCE WITH IRON-FOLIC ACID SUPPLEMENTATION AMONG PREGNANT MOTHERS IN BULE HORA DISTRICT, SOUTHERN ETHIOPIA: UNMATCHED CASE-CONTROL STUDY

**DOI:** 10.1101/2024.09.29.24314574

**Authors:** Reta Tesfa, Moges Mareg, Mahlet Birane, Midhagsaa Dhinsa, Biranu Muleta, Jibril Jemal, Tadase Begna

**Author notes:** Authors email address Moges Mareg ( /) Mahlet Birane Midhagsaa dhinsa Biranu Muleta Jibril Jemal Tadase Begna. Corresponding author: Reta Tesfa Department of Nursing, College of Health and Medical Sciences, Dilla University, Dilla, Ethiopia /+251961482912 Dilla P.O. Box: 419.

## Abstract

**Background:** World Health Organization has recommended a daily supplementation of iron folic acid, to avert anemias during pregnancy. However, due to many factors, compliance of pregnant women with this recommendation of iron and folic acid supplementation remains low, both in Africa and Ethiopia. The findings from previous studies show in concurrent finding regarding determinants of iron and folic acid compliance.

**Objective:** This study aimed to assess determinants of iron and folic acid supplementation compliance among antenatal care attendee pregnant women in the Bule Hora district, south Ethiopia, in 2023.

**Methods:** Health facility -based, unmatched case-control study was conducted in Bule Hora district from November 2023 to December 2023, using pretested interviewer-administered questionnaire. The sample size for the study was calculated using Epi Info version 7.2.6 software. A total of 115 cases and 230 controls were included in the study. The sample size was proportionally allocated to each health facility based on number of pregnant women supplemented with iron-folic acid at least one month before the data collection period; after that, systematic sampling techniques were used to select every 2^nd^ participant from each health facility. Binary and multivariable logistic regression was conducted to identify determinants of iron folic acid compliance; AOR at P-value < 0.05 with 95% CI was used to declare a statistically association after checking absence of multicollinearity (VIF < 1.65, Tolerance >0.6) and Hosmer and Leme-show test of model fitness (p-value =0.08).

**Results:** A total of 345 pregnant women were included in the study with, a 100% response rate. Prim gravidity [AOR: 4.67, 95% CI (1.60, 13.57)], antenatal care contact 4 or more times [AOR: 7.84, 95% CI (3.34-18.41)], having husband/family support to take iron folic acid [AOR: 4.48, 95% CI (2.19-9.13)] and good knowledge on anemia [AOR: 3.79, 95%CI (1.85-7.75)] shows significant association with iron-folic acid good compliance.

**Conclusion:** The study concluded that prim-gravidity, antenatal care contact 4 or more times, having husband support, and good knowledge of anemias were determinants of good compliance. Promoting husband support and frequency of antenatal care contact and providing information about anemias were needed to enhance compliance with iron folic acid.

## Introduction

Compliance with iron-folic acid supplements refers to how well patients follow their healthcare provider’s prescriptions regarding the specified dosage and timing of the supplements, which is usually measured as the number of times pregnant women take the supplements in a week [1, 2]. Iron folic acid supplementation is a provision of a formulation containing 60 mg of elemental iron and 400 micromilligrams of folic acid, especially during the pregnancy period of a woman’s life. The requirement for iron and folic acid is enhanced at the time of pregnancy due to hormonal and physiological changes during pregnancy. Poor compliance with the prescription of micronutrients during this period increases the likelihood of folic acid and iron deficiency [3, 4].

Deficiency of iron and folic acid is a worldwide micronutrient nutritional public health problem among women who are pregnant [5]. This iron and folic acid micronutrient deficiency leads to anemias in pregnant women [6].

Iron folic acid supplementation, iron-fortified staple foods, health and nutrition education, parasite infection control, and enhanced sanitation are the key practical and economical strategies for preventing and treating iron folic acid deficiency anemia in women who are pregnant [7]. As part of regular antenatal care, the WHO suggested initiation as early as possible and continuing a daily oral dose of 400μg folic acid and 60 mg iron supplements throughout pregnancy. Taking a recommended iron and folic acid supplements throughout pregnancy decreases the risk of iron deficiency anemia by 57% and the risk of all forms of maternal anemia by 70% at term [5].

Despite the role of daily IFAS in reducing anemia, compliance with IFAS remains moderately low in different parts of Africa, including Ethiopia [8-10]. Ethiopia set a significant aim to ensure that by 2029, 90% of women who are pregnant will receive iron-folic acid supplements for more than 90 days, according to the Ethiopian National Nutrition Program [11]. However, various studies reveal that IFAS compliance is still unacceptably low with rates varying from 92% in the Somali region to 50% in Addis Ababa [12]. This shows that there was setting specific variation in the level of compliance with IFAS among women who are pregnant, which mainly differs between rural and urban areas of the country. This is one of the main factors for IFAS programs failure as many experts believe on the issues [13]. If compliance is the main barrier to addressing IFA deficiency anemia, ways of improving it must be found.

Review of different literature on determinants of compliance with IFAS shows inconsistent findings [14, 15], and other equally important factors like media exposure and husband support have been given little attention. The low compliance in the current study area and the noted discrepancies in findings of previous studies on determinants of IFAS compliance and little emphasized factors, highlight the necessity of researching the variables affecting iron-folate supplement compliance and a thorough understanding of determinants of compliance with IFAS intake during pregnancy. To fulfill these gaps the current study used an unmatched case-control study design to identify determinants of compliance with IFAS among ante natal attendee pregnant women in the Bule Hora district, Oromia region, south Ethiopia.

## Method and Materials

### Study Area and Period

The study was conducted in Bule Hora district, south Ethiopia. Bule Hora district is found in the West Guji zone, Oromia region, and 472 km away from capital city, Addis Ababa, Ethiopia. The 2023 estimated population of the Bule Hora district is 292,701. The estimated number of pregnant women in the district is 10157. There are four health centers, 40 community health posts, and 26 private clinics providing maternal and child health services in the district. The study was conducted from November 2023 to December 2023.

### Study design

Health facility-based, unmatched case-control study was employed.

### Population

**Source population:** -All pregnant women who were attending antenatal care and who received iron and folate supplementation.

#### Study population

**Cases:** All pregnant women who were attending antenatal care and who took IFAS for less than 4 days per week in the recent week before the data collection period

**Control:** All pregnant women who were attending antenatal care and who took IFAS for at least 4 days per week in the recent week prior to the data collection period [8, 14, 16].

### Eligibility Criteria

#### Inclusion Criteria

All pregnant women who had antenatal care visits and supplemented with IFAS for a minimum of one month prior data collection period were included.

#### Exclusion Criteria

Pregnant women who were incapable of responding to the question (i.e., could not hear or talk) were excluded from the study.

### Sample size determination

The sample size of the study was calculated using the EPI Info program version 7.26, using two population proportion formulas for the determinants of IFAS compliance and corresponding parameters obtained from a recent case-control study done in Addis Ababa Ethiopia (46); by considering the following assumption: a case to control ratio of 1:2, 80% power and 95% confidence level and AOR=2.03(Table 1)

**Table 1:** Sample size calculation for a study on determinants of IFAS compliance among pregnant women attending ANC service in Bule Hora district, South Ethiopia 2023.

| S. n | Variable | CI % | Power % | case to control ratio | % of exposure among control | AOR | Calculated sample size |  |  | Reference |
| --- | --- | --- | --- | --- | --- | --- | --- | --- | --- | --- |
|  |  |  |  |  |  |  | Control | Cases | Total |  |
| 1 | ANC frequency | 95 | 80 | 1:2 | 73.1 | 4.21 | 84 | 42 | 126 | [17] |
| 2 | Gravidity | 95 | 80 | 1:2 | 35 | 2.03 | 209 | 105 | 314 | (46) |
| 3 | Knowledge on anemia | 95 | 80 | 1:2 | 30 | 2.3 | 79 | 158 | 237 | (14) |

From this, the study with the main factor, gravidity, gave a sufficiently large sample size (314), with a percentage of exposed (prim-gravida) among control (poor compliance) = 35% and a percentage of exposed among cases (good compliance) = 65.5% and an AOR of 2.03. As a result, 345 (115 cases and 230 controls) was used to conduct the study, after considering a 10% non-response rate.

### Sampling technique and procedure

Based on the sample size required for the study, a proportionate allocation of sample size was done for each health center based on the number of ANC case follow-ups and the total number of pregnant women in all health centers from the previous month’s ANC registration logbook. A systematic sampling technique was employed to select participants. Sample interval (K) was calculated by dividing the number of all pregnant women who took IFAS in the previous month from four health facilities in Bule Hora district which was 731, to the sample size needed to conduct the study which was 345, and the obtained result was 2. The first individual was selected by lottery method, and then every 2nd of the selected individuals was interviewed from each health center based on the allocated sample size until the required sample size was met. Accordingly, 84 participants from Bardaye Health Center, 126 from Garba Health Center, 96 from Killenso Mokonisa HC, and 39 from Killenso Rasa HC were participated in the study. (Fig: 1)

**Fig 1:**
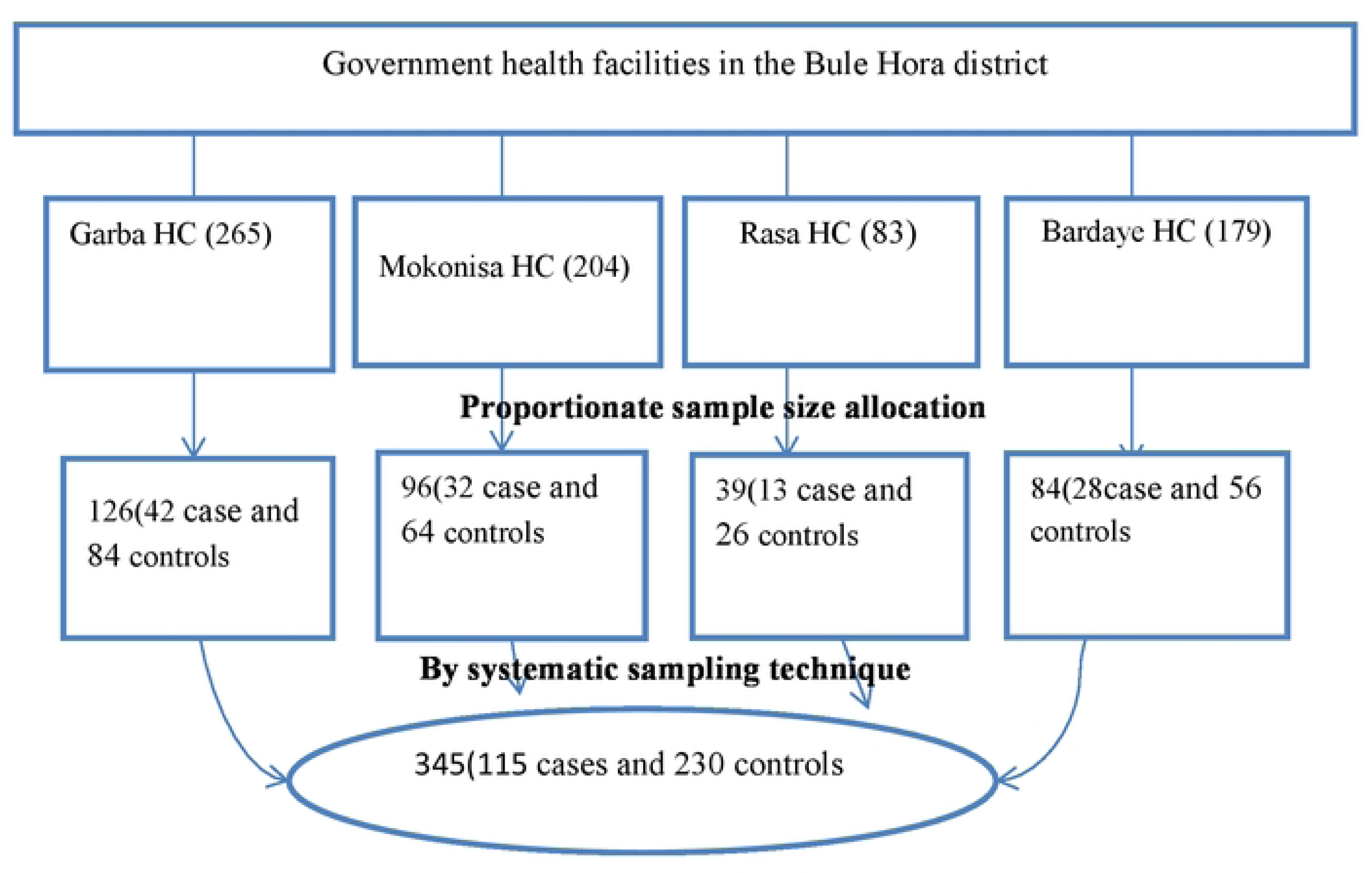
Schematic representation of sampling procedure to get study participants from Bule Hora district, public health facilities, west Guji zone, south Ethiopia, 2023

### Study Variables

#### Dependent variable

Compliance with iron-folic acid supplement

#### Independent variables

Socio-demographic related factors: age, marital status, residence area, father’s education level, family size, mother’s occupation, and mother’s education level.

Obstetric and non-obstetrics-related factors: ANC registration time, gravidity, and number of ANC visits, history of stillbirth and abortion, current anemia history.

Health service-related factors: Availability of IFAS, distance of home from the health facility, and counseling about iron folic acid during ANC service

Client-related factors: knowledge about Anemia and IFAS, media exposure, attitude towards IFAS and anemia, family/husband support

### Data collection tools and procedure

Data was collected by pretested interviewer-administered questionnaires and review of patient cards (to confirm the presence of current anemia). The questionnaires were designed after a review of the various literatures [2, 13, 17-24]. The questionnaire contains a total of 45 items, of which seven items were related to socio-demographic characteristics, 7 items were related to obstetrics and health, 4 items were related to health services, and 27 items were client-related factors. The midwife health professional in each health center determined the case status of the study participants, who were afterward linked to the data collectors with a unique identification code. A face-to-face interview technique was used to collect data by four clinical nurses under the close supervision of designated supervisors (2 BSc Nurses).

### Data quality management

The quality of data was assured by translating the English version of the pre-tested questionnaire to the local language, Afan Oromo, for interviews and then retranslated back to English for consistency. Two days of orientation were given for supervisors and data collectors on the study purpose and the data collection methods used, by the principal investigator. A week before the actual collection of data pretest of the questionnaire was done by using a 5% sample size of the population (17 participants) in Gwangua health center outside of the study area in the Abaya district. And, internal consistency of the questionnaire was assessed and Cronbach’s Alpha was computed; knowledge about IFAS (0.78), knowledge of pregnant women regarding anemia (0.842), and attitudes towards anemia and IFAS (0.868) which was acceptable. The data collectors were blinded from knowing the case status of the study participant. The principal investigator oversaw the entire process of data collection.

### Data Analysis Procedure

Data was entered into EPI Data version 4.1, then exported SPSS version 26. Before analysis, data completeness and missing value was checked. Descriptive statistics was computed to summarize categorical and continuous variables. Bi-variable analyses were conducted to identify candidate variables at p < 0.25, and those variables were further entered into a multivariable logistic regression model, to identify determinants of IFAS compliance among pregnant women.

Finally, p-values less than 5% with AOR at a 95% confidence level were used to declare variables with statistical significant association. Tolerance and the variance inflation factor (VIF) were used to test multicollinearity and determine if there was a linear association between the independent variables. All the variables were retained in multivariable analyses and none of them yielded variance inflation factors greater than 10, tolerance less than 0.1; (VIF<1.65, tolerance >0.6). Hosmer and Lemeshow’s test was found to be insignificant (p-value= 0.087). Depending on the kind of data collected, the study’s results were presented using tables, graphs, and text.

### Operational definition

Compliance with IFAS: Pregnant women who took the iron-folic acid supplement at least four days a week in the week before the data collection period. [1, 2, 14, 18, 24]

Good knowledge about iron folic acid supplementation: A questionnaire consisting 8 closed-ended questions was used to assess Women’s knowledge of IFAS (see annex-I, part III). Those who answered yes to the questions received a score of 1, and those who answered no to the questions received a score of 0. When a woman answered greater or equal to mean score of composite items, her knowledge of IFAS was rated as good; if she answered less than the mean score of composite items, her knowledge was rated as poor [24].

Good knowledge about anemia: a questionnaire consisting 6 closed-ended questions was used to measure the knowledge of pregnant women about anemia (see annex-I, part IV). Those who answered yes to the questions received a score of 1, and those who answered no to the questions received a score of 0. The woman’s knowledge regarding anemia was categorized as good for those who answered greater or equal to mean score of composite items and poor for those who answered below mean scores of composite items [18].

Attitude towards anemia and iron folate supplementation: - Ten closed-ended questions on anemia and IFA were posed to the respondents, and their responses were utilized to assess their attitude (see annex-I, part VI). A 5-point Likert scale was used to rate each of these items: 1 for “strongly agree,” 2 for “agree,” 3 for “not sure,” 4 for “disagree,” and 5 for “strongly disagree.” Based on the average of the ten items, a composite score was determined. Two categories were created from the composite score: below mean = positive attitude and, mean and above = negative attitude towards IFAS and anemia [25].

Current history of anemia: hemoglobin level less than 11mg/dl (confirmed by client card review) Media exposure: There were two categories for media exposure: satisfactory and unsatisfactory. Mothers were deemed to have satisfactory media exposure if they watched television or listened to the radio at least once a week; otherwise, they were deemed to have unsatisfactory media exposure [26].

### Ethical Consideration

The Helsinki Declaration was followed in the conduct of this study. Before data collection, ethical clearance was obtained from the institutional review board of Dilla University’s College of Health Science and Medicine (Ref. No: duchm/IRB/026/2023). A support letter was received from Dilla University’s Department of Human Nutrition. A permission letter was obtained from Bule Hora Health Office before data collection. Informed consent was received from participant and included information about the study’s objective and methods, possible risks and benefits, voluntary participation, and withdrawal rights. Attempt was done to keep all respondent data confidential. Respondents were also informed that their answers to the questions were grouped with other respondents’ answers and reported as part of a research study finding. No other ethical issue is related to the study.

## Result

### Socio-demographic characteristics of study participants

A total of 345 pregnant women (115 cases and 230 controls) were participated in this study, making the response rate of 100%. The mean (SD) age of study participants was 27.25 (±5.2) for cases and 26.7(±5.1) for controls. One hundred eighty-four (53.3%) of the pregnant women who participated in the study were found between the ages of 25-34 years, of whom 124(53.9%) were controls and 60(52.2%) were cases. From all participants majority of 324 (93.6%) of the pregnant women who participated were married and more than half of 202(58.6%) of them were housewives. Regarding their educational status only 72 (20.9%) of them cannot read and write. (Table 2)

**Table 2:**
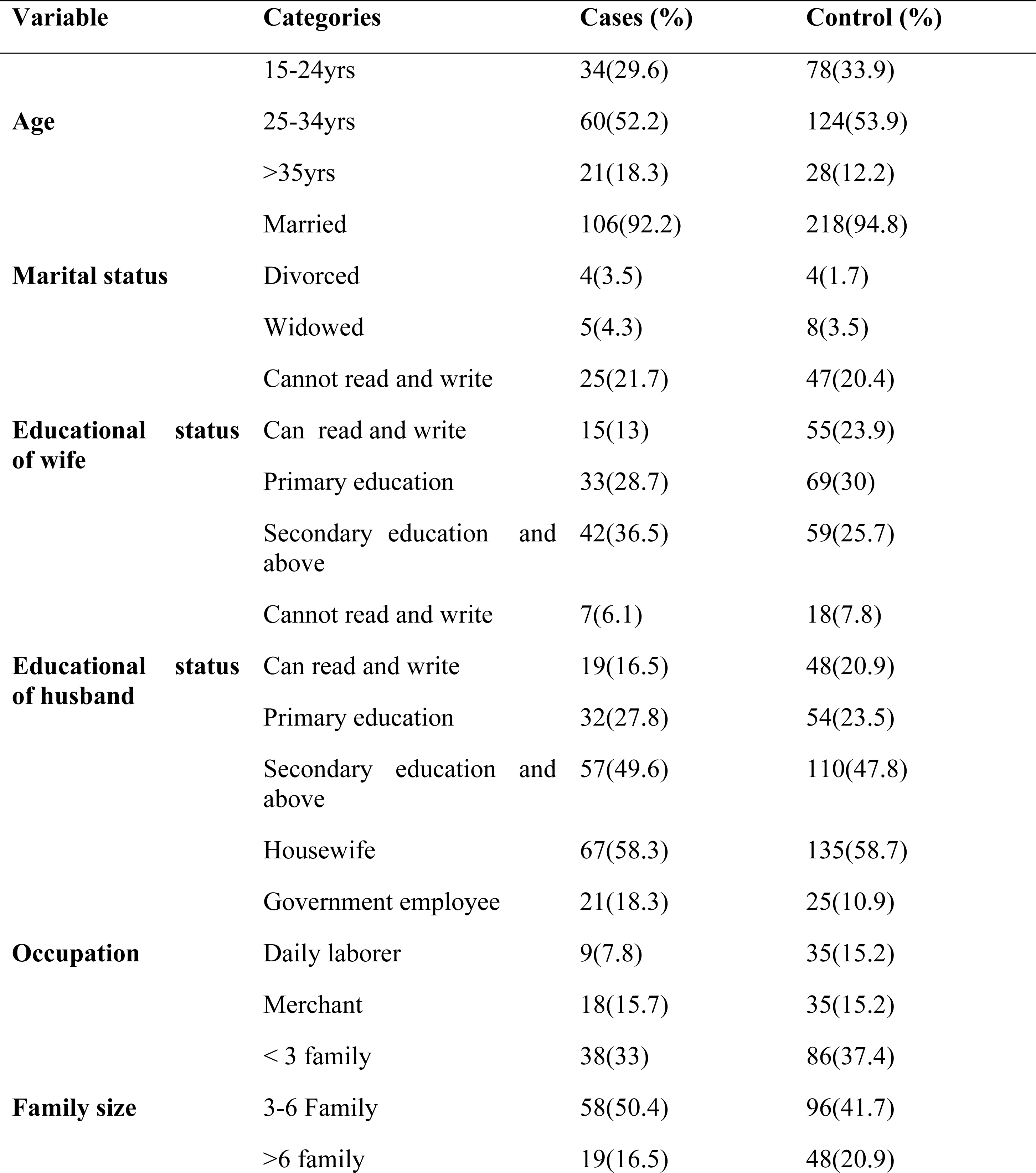
Socio-demographic characteristics of pregnant women attending ANC service ANC Bule Hora district health facilities, south Ethiopia 2023(n=345)

### Obstetric and non-obstetrics-related characteristics

Among the participants of the study, 106(92.2%) of cases and 193(83.9%) were multigravida mothers. Of the total participants, 14(4.1%) and 19(19.5%) have a history of stillbirth and a history of abortion respectively. Regarding ANC service 66(57.4%) cases and 163(709%) controls started ANC follow-up at the time of less than 16 week of their pregnancy. From 114(33%) of the participants who have four or more times ANC follow up during current pregnancy 8(7%) of them were cases and 106(46.1%) of them were controls. Regarding non obstetrics-related factors, 14(4.1%) and 19(5.5%) of pregnant mothers have a history of stillbirth and a history of abortion in their lifetime. (Table 3)

**Table 3:** Obstetric-related characteristics of pregnant women attending ANC service in Bule Hora district health facility, South Ethiopia 2023(n=345)

| Variable | Categories | Cases (%) | Control (%) | Total |
| --- | --- | --- | --- | --- |
| **Time of ANC visit** | ≤4month | 66(57.4) | 163(70.9) | 229(66.4%) |
|  | ≥4month | 49(42.6) | 67(29.1) | 116(33.6%) |
| **Frequency of ANC follow-up** | Less than 4 | 107(93) | 124(53.9) | 231(67%) |
|  | 4 or more times | 8(7) | 106(46.1) | 114(33%) |
| **Gravidity** | Multigravida | 106(92.2) | 193(83.9) | 299(86.7%) |
|  | prim gravida | 9(7.8) | 37(16.1) | 46(13.3%) |
| **Having a current history of anemia** | No | 84(73) | 185(80.4) | 269(78%) |
|  | Yes | 31(27) | 45(19.6) | 76(22%) |
| **Ever had a history of abortion** | No | 108(93.9) | 218(94.8) | 326(94.5%) |
|  | Yes | 7(6.1) | 12(5.2) | 19(5.5%) |

### Health service-related factors

Regarding distance from health facilities, 200(58%) of pregnant mothers travel less than 30 minutes to reach health facilities which accounts 70(35%) of cases and 130(65%) of controls, but the remaining 35(10.1%) of them travel more than one hour from which 12(34.3%) were cases and 23(65.7) were controls. Waiting time at health facilities was between 15 to 30 minutes for the majority of the respondents which accounts 81(70.4%) of cases and 169(73.5) of controls. Regarding IFAS, 268(77.7%) of pregnant women obtain adequate supplements from health facilities, from which 72(62.6%) of them were cases and 196(85.2%) of them were controls. Around 258(74.8%), of the participants got counseling from service providers about the supplementation, from which 69(60%) of them were cases and 189(82.2%) of them were controls.

### Client related factors

Out of the total participants, 186(53.9%) have access to media (radio/television) of which 61(53%) were cases and 125(54.3%) were controls. Pregnant women who reported husband/family support to take IFA tablet accounts 175 (50.7%) of the respondents, among them 31(27%) were cases and 144(62.6%) were controls. (Fig: 2)

**Fig 2:**
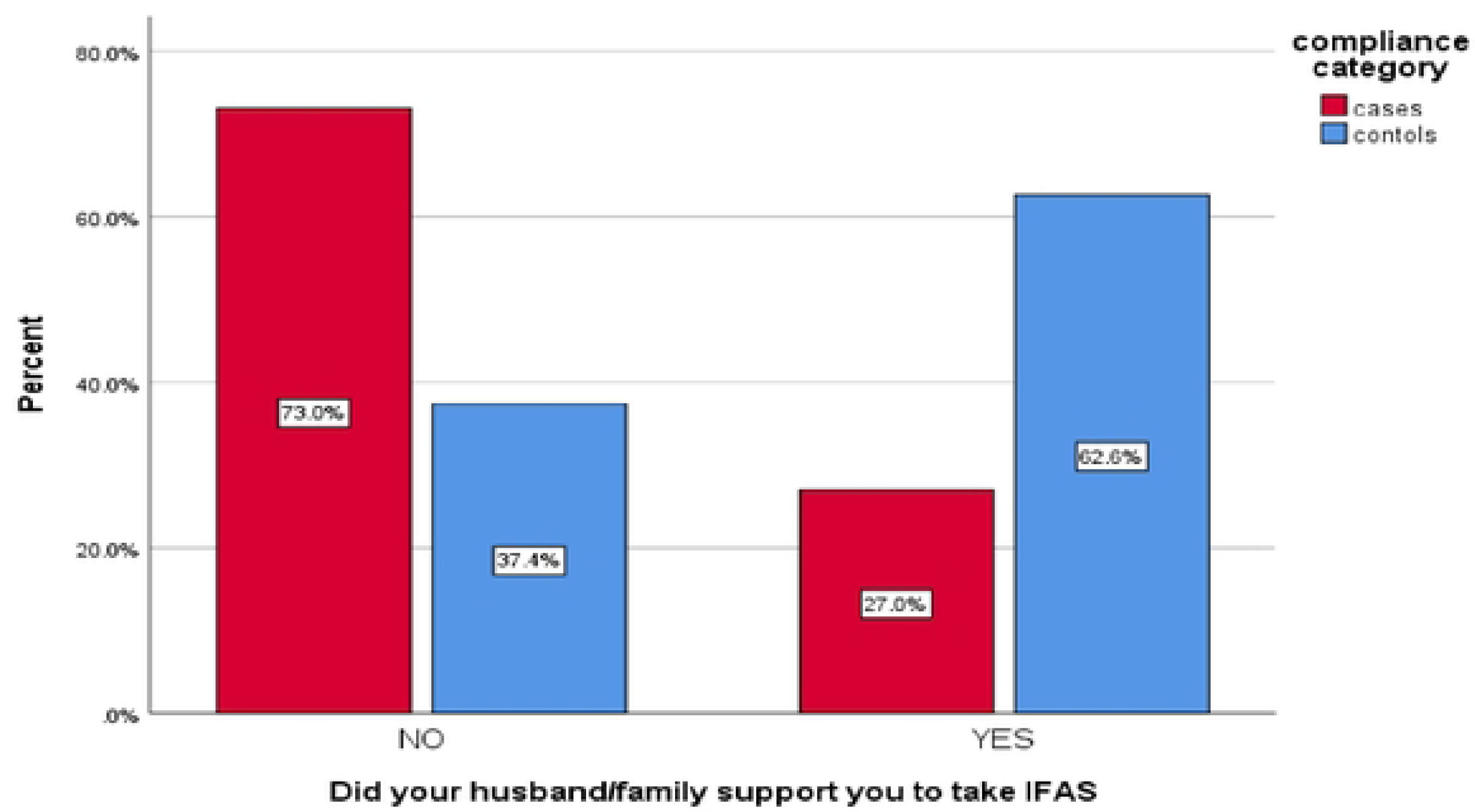
Number of pregnant women who have husband support to take IFAS, 1n Buie Hora district health facility, south Ethiopia 2023(n=345)

### Knowledge of pregnant women on IFAS and anemia

Of all participants, 88 (29.7%) cases and 208 (70.3%) controls knew the benefits of taking IFAS tablets during pregnancy. Only 25 (28.1%) of cases and 64 (71.9%) of controls knew importance of taking iron and folic acid tablets in postpartum period. Participants 88(29.7%) of cases and 208(70.3%) of controls knew that taking iron and folic acid supplement during pregnancy is crucial to the mother. Only 25(28.1%) of cases and 64(71.9%) of controls knew about the health benefit of continuing iron and folic acid tablets in the postpartum period. (Table 4)

**Table 4:** Response of pregnant women attending ANC service, on IFAS-related questions, in Bule Hora district health facility, south Ethiopia 2023 (n=345)

| s.no | Question | Response | Cases (%) | Controls (%) |
| --- | --- | --- | --- | --- |
| 1 | Have you ever heard about IFAS? | No | 33(45.2%) | 40(54.8%) |
|  |  | Yes | 82(30.1%) | 190(69.9%) |
| 2 | Taking iron folic acid during pregnancy is crucial for the mother | No | 27(55.1%) | 22(44.9%) |
|  |  | Yes | 88(29.7%) | 208(70.3%) |
| 3 | Taking iron folic acid during pregnancy is crucial for the fetus | No | 47(45.6%) | 56(54.4%) |
|  |  | Yes | 68(28.1%) | 174(71.9%) |
| 4 | Taking iron folic acid begins from confirmation of pregnancy and continue | No | 88(39.3%) | 136(60.7%) |
|  |  | Yes | 27(22.3%) | 94(77.7%) |
| 5 | Taking iron folic acid during pregnancy is crucial to avoid anemia | No | 50(60.2%) | 33(39.8%) |
|  |  | Yes | 65(24.8%) | 197(75.2%) |
| 6 | Taking IFA tablets continues during the postpartum period | No | 90(35.2%) | 166(64.8%) |
|  |  | Yes | 25(28.1%) | 64(71.9%) |
| 7 | Consuming IFA tablets during pregnancy does not lead to a big baby | No | 62(47.7%) | 68(52.3%) |
|  |  | Yes | 53(24.7%) | 162(75.3%) |
| 8 | Taking IFA pills during pregnancy may help prevent birth abnormality | No | 63(45.7%) | 75(54.3%) |
|  |  | Yes | 52(25.1%) | 155(74.9%) |

Regarding responses of the participants to the questions asked to assess their knowledge of anemia 74(25.3%) of cases and 219(74.6%) of controls knew that anemia can be prevented. Among all respondents, only 22(21.2%) of cases and 82(78.8%) of controls knew about anemic women becoming breathless easily. (Table 5)

**Table 5:** Response of pregnant women attending ANC service, on anemia-related questions, in Bule Hora district health facility, south Ethiopia 2023 (n=345)

| s.no | Question | Response | Cases (%) | Controls (%) |
| --- | --- | --- | --- | --- |
| 1 | Pregnancy can make women anemic | No | 65(42.8%) | 87(57.2%) |
|  |  | Yes | 50(25.9%) | 143(74.1%) |
| 2 | Anemic women become breathless easily | No | 93(38.6%) | 148(61.4%) |
|  |  | Yes | 22(21.2%) | 82(78.8%) |
| 3 | Anemic women have weaknesses | No | 56(56.6%) | 43(43.4%) |
|  |  | Yes | 59(24.0%) | 187(76.0%) |
| 4 | Anemic women have pale skin or tongue | No | 88(36.2%) | 155(63.8%) |
|  |  | Yes | 27(26.5%) | 75(73.5%) |
| 5 | Anemia can be prevented | No | 41(78.8%) | 11(21.2%) |
|  |  | Yes | 74(25.3%) | 219(74.7%) |
| 6 | Are you aware of the measures taken to prevent anemia? | No | 43(70.5%) | 18(29.5%) |
|  |  | Yes | 72(25.4%) | 212(74.6%) |

Regarding overall knowledge of the pregnant women about IFAS and anemia, 220(63.8%) and 183(53%) of them were categorized as having good knowledge of IFAS and good knowledge of anemia respectively. Among the women having good knowledge of IFAS 52(45.2%) of them were cases and 168(73%) of them were controls, and of women having good knowledge of anemia 35(30.4%) of them were cases, and 82(35.7%) of them were controls. (Table 6)

**Table 6:** knowledge of pregnant women attending ANC service, on IFAS and anemia, in Bule Hora district health facility, south Ethiopia 2023 (n=345)

| Variable | Categories | Cases (%) | Control (%) | Total (%) |
| --- | --- | --- | --- | --- |
| **Knowledge on IFAS** | Poor knowledge | 63(54.8) | 62(27) | 125(36.2) |
|  | Good knowledge | 52(45.2) | 168(73) | 220(63.8) |
| **Knowledge on anemia** | Poor knowledge | 80(69.6) | 82(35.7) | 162(47) |
|  | Good knowledge | 35(30.4) | 148(64.3) | 183(53) |

### The attitude of pregnant women toward IFAS and anemia

Regarding pregnant women’s attitudes towards IFAS and anemia 197(57.1%) of them were categorized as having positive among whom 45((39.13%) were cases and 152(66.09%) of them were controls. (Fig: 3)

**Fig 3.**
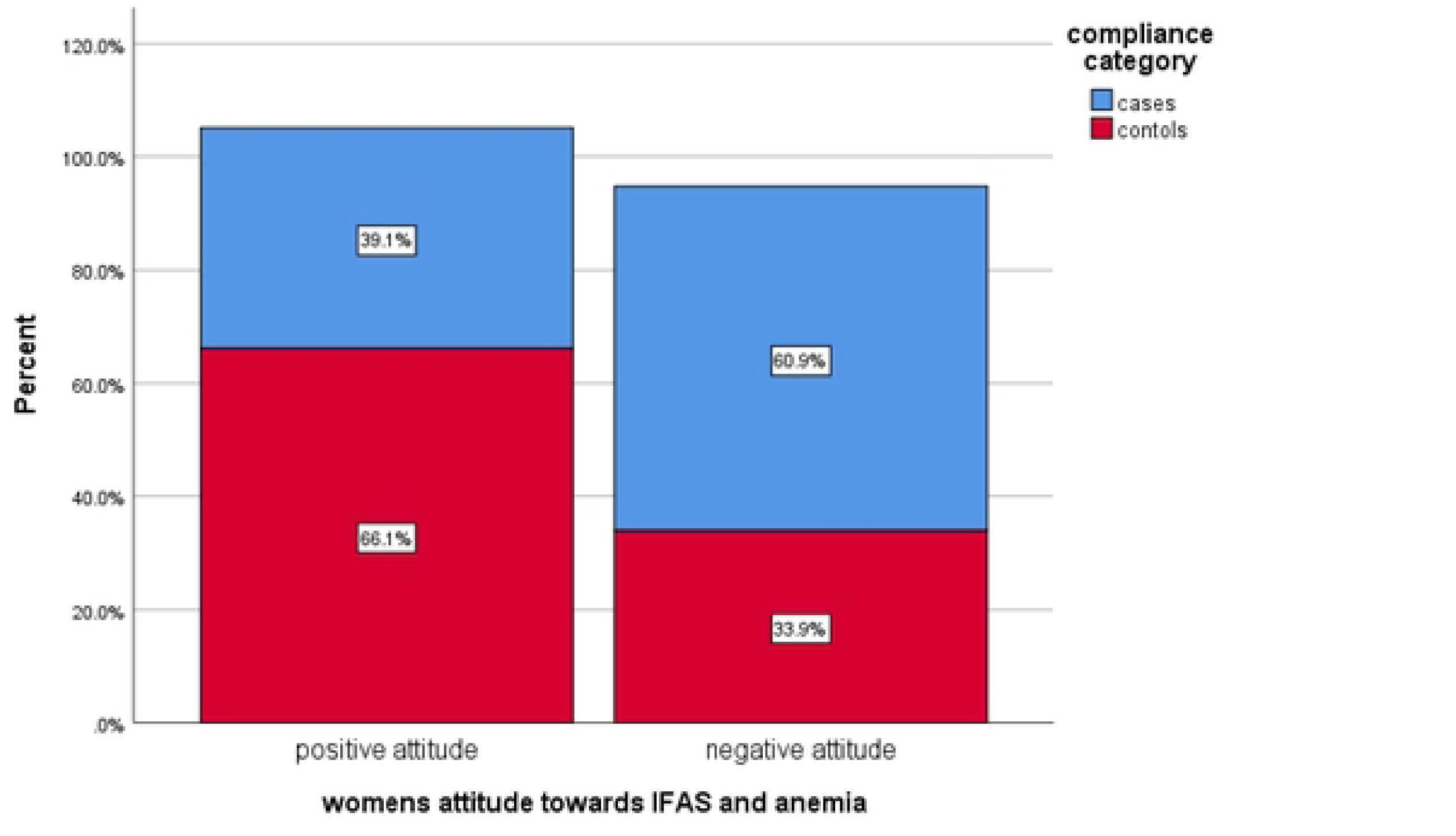
: Attitude of pregnant women attending ANC service, towards IFAS and anemia, in Bule Hora district health facility, south Ethiopia 2023(n=345)

### Determinants of compliance with IFAS among pregnant women

Bivariable analysis revealed that at a p-value less or equal to 0.25 significance level, 13 variables namely occupational status, educational status, residency, gravidity, frequency of ANC visit, time at ANC visit starts, current history of anemia, obtaining adequate IFAS tablet from health facilities, getting counseling from health provider about IFAS, husband/family support to take IFAS, knowledge on anemia and IFAS, and attitude toward IFAS and anemia were identified as; candidate variables for multivariable logistic analysis. All candidate variables were entered together into a multivariable logistic regression to determine the final determinants of IFAS compliance. In a multivariable logistic regression, four variables were retained statistical significant at p-value of < 0.05 and at 95% confidence level. Accordingly, gravidity, frequency of ANC visits; husband /family support, and knowledge of anemia were significantly associated with IFAS compliance

According to this study, the odds of having good compliance with iron and folic acid supplementation were 4.67 times higher among prim gravida women when compared to multigravida women. The study also revealed that the odds of having good compliance with IFAS were 7.84 times higher among pregnant women who have ANC visits 4 or more times when compared to those who have less than 4 times ANC visits during their current pregnancy. (Table 7)

**Table 7:** Bi-variable and multivariable logistic regression analysis of determinants of IFAS compliance among pregnant women attending ANC at Bule Hora district, South Ethiopia, 2023(n=345)

| Variable | Category | Cases (%) | Controls (%) | COR (95% CI) | AOR (95% CI) | p-value |
| --- | --- | --- | --- | --- | --- | --- |
| **Gravidity** | Multigravida | 106(92.17) | 193(83.91) | 1 | 1 |  |
|  | prim gravida | 9(7.83) | 37(16.09) | 2.25(1.05-4.8) | 4.67(1.60-13.57) | 0.005** |
| **Frequency of ANC visit** | Less than 4 | 107(93.04) | 124(53.91) | 1 | 1 |  |
|  | 4 or more times | 8(6.96) | 106(46.09) | 11.43(5.32-24.54) | 7.84(3.34-18.41) | <0.001** |
| **Husband /family support** | No | 84(73.04) | 86(37.39) | 1 | 1 |  |
|  | Yes | 31(26.96) | 144(62.61) | 4.53(2.77-7.41) | 4.48(2.19-9.13) | <0.001** |
| **Knowledge on anemia** | Poor knowledge | 80(69.57) | 82(35.65) | 1 | 1 |  |
|  | Good knowledge | 35(30.4) | 148(64.35) | 4.12(2.55-6.66) | 3.79(1.85-7.75) | <0.001** |
| **COR crude odds ratio and AOR adjusted odds ratio \*P ≤ 0.05, \*\*P<0.001 Hosmer and Leme-show test=0.087** |  |  |  |  |  |  |

Furthermore, women those having good knowledge of anemia were 3.79 times more likely to have compliance with iron and folic acid supplementation than those who had poor knowledge by keeping other variables constant. (Table 7)

The study also shows that the odds of having good compliance with iron and folic acid supplementation were 4.8 times higher among those who have husband support when compared to those who have no husband support. (Table 7)

## Discussion

The World Health Organization has recommended daily iron and folic acid supplementation starting in the first trimester to avert anemia during pregnancy. Ethiopia also incorporates this recommendation into a free antenatal program. However, due to many factors, compliance of pregnant women with this recommendation of iron and folic acid supplementation remains low, both in Africa and Ethiopia. This study was tried assess determinants of iron folic acid compliance among pregnant women attending ANC service in Bule Hora district.

This study revealed that being prim gravida mothers were positively associated with IFAS compliance. The finding was in line with cross-sectional studies done on adherence to iron and folic acid supplementation during pregnancy in Kenya and in western Uganda [8, 10, 27]. But this finding was different from the finding of studies done at Ayder Comprehensive Specialized Hospital, at Debre Tabor General Hospital in northern Ethiopia, and Asela town, Oromia region, which indicate that multi gravida mothers were more likely to have compliance with IFAS than prim gravida [21, 28, 29]. The possible explanation for the current finding might be that prim gravida mothers lack birth experience; they perceive a higher level of pregnancy-related risk than multigravida mothers do, which may explain why they are more likely to follow health care provider recommendations of IFAS intake.

The other determinant identified in this study was the frequency of ANC visits. In this study, pregnant mothers who have ≥4 ANC visits were more likely to have compliance with IFAS than pregnant women who have ANC visits less than four. This finding is in line with the study conducted in the western zone of Tigray, Assela town, Debre Tabor General Hospital, and Public hospitals of Dire Dawa [17, 21, 29, 30]. The possible explanation could be that pregnant women who had more ANC visits were more likely to receive more ANC services, including counseling, and more likely to acquire better information and knowledge about the health benefit of iron and folic acid to prevent anemia during pregnancy. This gives the healthcare professional an excellent chance to address any issues pregnant women may have while taking the supplements and to encourage the mothers to utilize them as prescribed.

The study also revealed that pregnant women who had good knowledge about anemia were more likely to have compliance with iron and folic acid supplementation than their counterparts. This study was in line with a study conducted in Adwa town Tigray region, Northern Ethiopia. This finding was also supported by a study done in the Dembia district and the North Wollo zone of Northern Ethiopia [20, 22]. This result could be explained by the idea that a mother who knows more about anemia will be able to comprehend its causes, preventative strategies, and potential problems that could harm both the mother and her unborn child. The mothers learn from this how important it is to take IFA as prescribed.

In this study, pregnant mothers whose husbands/families supported them to take IFAS were more likely to comply with IFAS than those who did not have their husbands’ support. This finding was supported by the study done in Adwa town Tigray region Northern Ethiopia [30]. This might be because the husband is the closest person to his wife so he helps remind her to take her IFA tablets daily.

In conclusion, the finding of the current study shows that pregnant women who did not have compliance with IFAS were those who were multi-gravida, who had ANC contact less than four times during pregnancy, had low knowledge of the risk of anemia, and who had a negative attitude towards IFAS and anemia.

### Limitation of the study

Pregnant women’s self-reports provided information on the determinants of compliance with iron-folic acid, which could lead to recall bias and social desirability bias because respondents might report what was expected of them while their actual compliance with IFAS intake might differ. This could lead to misclassification bias when determining study participants’ compliance status.

### Strength of the study

The strength of the study was that it was a multicentered study and involved all health facilities in Bule Hora district, which increased its generalizability. Moreover, the study tried to address many determinants of IFAS compliance and provided good information for an intervention plan to enhance compliance with IFAS among pregnant mothers.

## Conclusion

In the current study, we found that being prim gravida, having ANC visits 4 and more, having good knowledge of anemia, and having husband support to remind them to take IFAS were factors that positively influenced pregnant women compliance with iron and folic acid supplementation.

### Recommendation

Based on the findings of this study, the following recommendations were forwarded:-

#### For health facilities

➢ Encouraging health professionals to provide health education to increase awareness about anemia consequences during pregnancy
➢ Giving attention to multigravida mother while giving ANC service
➢ Encouragement of male involvement in ANC follow-up.

#### For the District health office

➢ Raising community awareness on iron folate supplementation benefits and the danger of anemia for pregnant women through health education at regular community meetings and mass media campaigns is highly recommended.
➢ Moreover, trying to boost ANC visits by providing little incentives such as a maternal welcoming package during ANC visits and issuing a certificate to mothers who complete more than four ANC visits could enhance IFAS compliance.

#### For Researchers

➢ Further studies supplemented with in-depth qualitative studies to address sociocultural determinants, through objective compliance measurement methods such as the pill count method are recommended

## Declarations

### Ethics approval and consent to participate

The Helsinki Declaration was followed in the conduct of this study. Before data collection, ethical clearance was obtained from the institutional review board of Dilla University’s College of Health Science and Medicine (Ref. No: duchm/IRB/026/2023). A support letter was received from Dilla University’s Department of Human Nutrition. A permission letter was obtained from Bule Hora Health Office before data collection. Informed consent was received from participant and included information about the study’s objective and methods, possible risks and benefits, voluntary participation, and withdrawal rights. Attempt was done to keep all respondent data confidential. Respondents were also informed that their answers to the questions were grouped with other respondents’ answers and reported as part of a research study finding. No other ethical issue is related to the study.

### Consent for publication

Not applicable

### Availability of data and materials

Upon a reasonable request, the corresponding author will provide all study data.

### Author contributions

All authors have substantial contribution on the conception, design analysis and interpretation of results. All authors read and approved the final manuscript.

## Data Availability

All relevant data are within the manuscript and its supporting information files.

## Acknowledgement

I would like to acknowledge Dilla University, college of health science and medicine, Department of Human Nutrition, for providing me opportunity conduct this Master’s thesis. Additionally, I want to express my heartfelt gratitude and appreciation to my advisors, Mr. Moges Mareg (MPH, Assistant professor) and Mrs. Mahlet Birhane (BSc,MSc), for their advice, encouragement, insightful comments, and suggestions throughout the course of my study.

I would also like to express my gratitude to the staff of Bule Hora district health office, data collectors, and supervisors for their cooperation and invaluable support during data collection. I want to express my appreciation to the study subject for their participation. Finally, I would like to extend my special thanks to my friends and family members for their dedicated support to finalize my study.

## References

1. Brown, M.T. and J.K. Bussell, Medication Adherence: WHO Cares? Mayo Clinic Proceedings, 2011. 86(4): p. 304–314.

2. Tegodan, E., G. Tura, and A. Kebede, Adherence to Iron and Folic Acid Supplements and Associated Factors Among Pregnant Mothers Attending ANC at Gulele Sub-City Government Health Centers in Addis Ababa, Ethiopia. Patient Prefer Adherence, 2021. 15: p. 1397–1405.

3. Habib, F., et al., Compliance to iron supplementation during pregnancy. Journal of Obstetrics and Gynaecology, 2018. 29(6): p. 487–492.

4. Organization, W.H., WHO recommendations on antenatal care for a positive pregnancy experience. 2016: World Health Organization.

5. Organization, W.H., Guideline: daily iron and folic acid supplementation in pregnant women. 2012: World Health Organization.

6. Federal Ministry of Health - FMoH, National Antenatal Care Guideline. 2022, Addis Ababa: Federal Ministry of Health - FMoH,.

7. Berti, C., M. Faber, and C.M. Smuts, Prevention and control of micronutrient deficiencies in developing countries: current perspectives. Nutrition and Dietary Supplements, 2014. 6: p. 41–57.

8. Nimwesiga, C., M. Murezi, and I.M. Taremwa, Adherence to Iron and Folic Acid Supplementation and Its Associated Factors among Pregnant Women Attending Antenatal Care at Bwindi Community Hospital, Western Uganda. Int J Reprod Med, 2021. 2021: p. 6632463.

9. Pathirathna, M.L., et al., Maternal Compliance to Recommended Iron and Folic Acid Supplementation in Pregnancy, Sri Lanka: A Hospital-Based Cross-Sectional Study. Nutrients, 2020. 12(11).

10. Bahati, F., S. Kairu-Wanyoike, and J.M. Nzioki, Adherence to iron and folic acid supplementation during pregnancy among postnatal mothers seeking maternal and child healthcare at Kakamega level 5 hospital in Kenya: a cross-sectional study. Wellcome Open Res, 2021. 6: p. 80.

11. Ayana, G., et al., Ethiopian National Nutrition Program End-Line Survey. 2015.

12. (MoH), T.E.P.H.I.E.i.c.w.t.U.E.a.t.M.o.H., Food and Nutrition Strategy (FNS) baseline survey. 2023: Addis Ababa,Ethiopia.

13. Ethiopian Public Health Institute - EPHI, Federal Ministry of Health - FMoH, and ICF, Ethiopia Mini Demographic and Health Survey 2019. 2021, EPHI/FMoH/ICF: Addis Ababa, Ethiopia.

14. MoH, National Iron and Folic Acid Supplementation; Communication Strategy, 2013-2017. Division of Nutrition, 2013.

15. Hyder, S.M., et al., Do side-effects reduce compliance to iron supplementation? A study of daily- and weekly-dose regimens in pregnancy. J Health Popul Nutr, 2002. 20(2): p. 175–9.

16. Taye, T.A., M. Sinaga, and A. Taye, Determinants of adherence to iron-folic acid supplementation among postnatal mothers in Addis Ababa referral hospitals, Ethiopia. 2021, Research Square.

17. Solomon, Y., A. Sema, and T. Menberu, Adherence and associated factors to iron and folic acid supplementation among pregnant women attending antenatal care in public hospitals of Dire Dawa, Eastern Ethiopia. European Journal of Midwifery, 2021. 5(August): p. 1–7.

18. Mamo, T.T., et al., Adherence to prenatal iron-folic acid supplementation and associated factors among pregnant women attending antenatal care services in Dilla town, South Ethiopia. Med Access Point Care, 2021. 5: p. 23992026211008805.

19. Seifu, C.N., S.J. Whiting, and T.G. Hailemariam, *Better-Educated,* Older, or Unmarried Pregnant Women Comply Less with Iron–Folic Acid Supplementation in Southern Ethiopia. Journal of Dietary Supplements, 2020. 17(4): p. 442–453.

20. Molla, T., et al., Factors associated with adherence to iron folate supplementation among pregnant women in West Dembia district, northwest Ethiopia: a cross sectional study. BMC Res Notes, 2019. 12(1): p. 6.

21. Gebremariam, A.D., et al., Adherence to iron with folic acid supplementation and its associated factors among pregnant women attending antenatal care follow up at Debre Tabor General Hospital, Ethiopia, 2017. PLoS One, 2019. 14(1): p. e0210086.

22. Demis, A., et al., Iron and folic acid supplementation adherence among pregnant women attending antenatal care in North Wollo Zone northern Ethiopia: institution based cross-sectional study. BMC Res Notes, 2019. 12(1): p. 107.

23. Siabani, S., et al., Determinants of Compliance With Iron and Folate Supplementation Among Pregnant Women in West Iran: A Population Based Cross-Sectional Study. J Family Reprod Health, 2018. 12(4): p. 197–203.

24. Boti, N., et al., Adherence to Iron-Folate Supplementation and Associated Factors among Pastoralist’s Pregnant Women in Burji Districts, Segen Area People’s Zone, Southern Ethiopia: Community-Based Cross-Sectional Study. Int J Reprod Med, 2018. 2018: p. 2365362.

25. Rai, S.S., et al., Effect of knowledge and perception on adherence to iron and folate supplementation during pregnancy in Kathmandu, Nepal. Journal of the Medical Association of Thailand = Chotmaihet thangphaet, 2019. 97 Suppl 10: p. S67–74.

26. Asmamaw, D.B., et al., Poor adherence to iron-folic acid supplementation and associated factors among pregnant women who had at least four antenatal care in Ethiopia. A community-based cross-sectional study. Front Nutr, 2022. 9: p. 1023046.

27. Kamau, M.W., W. Mirie, and S. Kimani, Compliance with Iron and folic acid supplementation (IFAS) and associated factors among pregnant women: results from a cross-sectional study in Kiambu County, Kenya. BMC Public Health, 2018. 18(1): p. 580.

28. Gebremichael, T.G., H. Haftu, and T.A. Gereziher, Time to start and adherence to iron-folate supplement for pregnant women in antenatal care follow up; Northern Ethiopia. Patient Prefer Adherence, 2019. 13: p. 1057–1063.

29. Murugan, R., Determinants of Adherence to Iron Folic Acid Supplementation among Pregnant Women Attending Antenatal Clinic in Asella Town, Ethiopia. 2018. 35.

30. Gebremichael, T.G. and T.G. Welesamuel, Adherence to iron-folic acid supplement and associated factors among antenatal care attending pregnant mothers in governmental health institutions of Adwa town, Tigray, Ethiopia: Cross-sectional study. PLoS One, 2020. 15(1): p. e0227090.

